# Microbial GWAS studies revealing combinations of Omicron RBD mutations existed and may contribute to antibody evasion and ACE2 binding

**DOI:** 10.1101/2022.01.19.22269510

**Authors:** Xumin Ou, Zhishuang Yang, Dekang Zhu, Sai Mao, Mingshu Wang, Renyong Jia, Shun Chen, Mafeng Liu, Qiao Yang, Ying Wu, Xinxin Zhao, Shaqiu Zhang, Juan Huang, Qun Gao, Yunya Liu, Ling Zhang, Maikel Peopplenbosch, Qiuwei Pan, Anchun Cheng

**Author notes:** Anchun Cheng. **Email:**. **Author Contributions:** X.O., Q.P. and A.C. devised the project and the main conceptual ideas; X.O., Z.Y., D.Z., S.M. and M.W. acquired data; X.O. and Z.Y. designed and performed the computations; X.O., Z.Y., D.Z., S.M., R.J., S.C., M.L. and Q.Y. analyzed and interpreted data; X.O. and Z.Y. drafted the manuscript; Y.W., X.Z., S.Z., J.H., Q.G., Y.L., L.Z., M.P. and Q.P. proofread the draft; all authors approved the final version of the manuscript.

## Abstract

Since Omicron variant of SARS-CoV-2 was first detected in South Africa (SA), it has now dominated in United Kingdom (UK) of Europe and United State (USA) of North America. A prominent feature of this variant is the gathering of spike protein mutations, in particularly at the receptor binding domain (RBD). These RBD mutations essentially contribute to antibody resistance of current immune approaches. During global spillover, combinations of RBD mutations may exist and synergistically contribute to antibody resistance in fact. Using three geographic-stratified genome wide association studies (GWAS), we observed that RBD combinations exhibited a geographic pattern and genetical associated, such as five common mutations in both UK and USA Omicron, six or two specific mutations in UK or USA Omicron. Although the UK specific RBD mutations can be further classified into two separated sub-groups of combination based on linkage disequilibrium analysis. Functional analysis indicated that the common RBD combinations (fold change, -11.59) alongside UK or USA specific mutations significantly reduced neutralization (fold change, -38.72, -18.11). As RBD overlaps with angiotensin converting enzyme 2(ACE2) binding motif, protein-protein contact analysis indicated that the common RBD mutations enhanced ACE2 binding accessibility and were further strengthened by UK or USA-specific RBD mutations. Spatiotemporal evolution analysis indicated that UK-specific RBD mutations largely contribute to global spillover. Collectively, we have provided genetic evidence of RBD combinations and estimated their effects on antibody evasion and ACE2 binding accessibility.

## Introduction

The Omicron variant of SARS-CoV-2 was recently detected in southern Africa, and has now sparked a new wave of global epidemic, in particular Europe and North America. A notable feature of this variant is the large amount of RBD mutations on spike protein (1). One of fundamental impacts of these mutations is antibody resistance (so called antibody evasion) (2). Because neutralization by clinically used monoclonal antibodies or serum of post COVID-19 vaccination is completely or incompletely diminished (3). This has been confirmed by evaluating neutralizing activity of 17 commercial antibodies to spike protein or pseudo-virus with differently individual RBD mutation (3). As a receptor docking domain of spike protein, these mutations may also change viral entry via regulating binding affinity to ACE2 receptor (4). Thus, the RBD mutations not only participate in antibody evasion but also ACE2 binding activity.

However, the RBD mutations may collaboratively contribute to antibody evasion, in a form of RBD combination or an epitope but not individual mutation. To address this question, we aimed to know if combinations of RBD mutations of Omicron variant exist and how they contribute to antibody evasion and ACE2 binding activity. In order to further understand the trend of its current global transmission, spatiotemporal evolution of Omicron harboring different RBD combinations will be studied.

## Results

### Different combinations of RBD mutations existed in United Kingdom and United state Omicron

As of 2-Jan-2022, we retrieved 77123 sequences of Omicron variant from GASID database. This variant was highly distributed in Africa, Europe and North America, in particular south Africa (SA) (72.94%, 1383/1896), United Kingdom (UK) (77.91%, 39849/51144) and United State (USA) (95.88%,19534/20374) (Fig.1A). To understand if geographic combinations of RBD mutations existed, three microbial-Genome wide association studies (GWASs) and linkage disequilibrium (LD) analysis were conducted between SA, UK and USA (5). Comparing with the SA Omicron, we observed that S477N, T478K, E484A, Q493R and G496S were significantly enriched in both USA and UK Omicron, and defined as common RBD mutations in this context (Fig.1B). However, the percentage of enrichment of these mutations in USA Omicron is much lower than that of UK Omicron. Meta-analysis of three GWAS studies showed that S371L, S373P, S375F, Q498R, N501Y and N505H were specifically enriched in UK Omicron, and G446S and N440K specifically in USA Omicron (Fig.1C-D). In addition, genetic association analysis indicated that common increased RBD mutations alongside with UK or USA specific RBD mutations were genetically linked or concurrently existed in Omicron variants (Fig.1E and Dataset).

**Figure 1.**
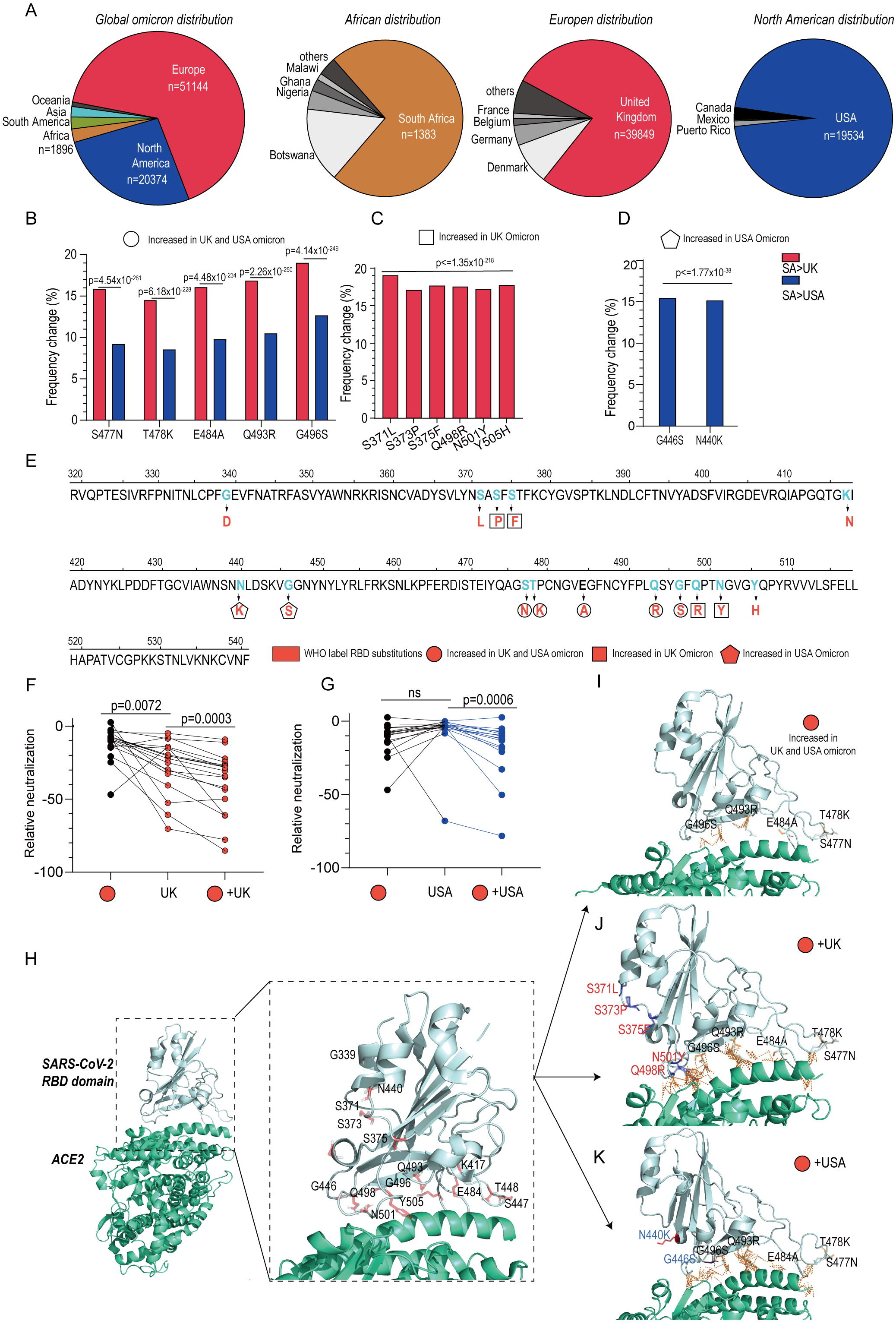
Combinations of RBD mutations and corresponding effects on antibody neutralization and ACE2 binding. (A) The South Africa (SA) isolates, United Kingdom (UK) isolates and USA isolates dominated the global population, as of 2-Jan-2022. (B) Commonly increased but frequency differently RBD mutations identified in UK and USA Omicron variants. Three geographical stratified GWAS studies (SA>UK, SA>USK and SA>USA) and Linkage Disequilibrium (LD) analysis were conducted. (C-D) UK and USA-specific RBD mutations. (E) WHO defined RBD substitutions and GWAS studies identified common, and specific combinations of RBD substitution in UK and USA Omicron. (F-G) Antibody resistance to 17 commercial mono-antibody was enhanced by combinations of RBD mutations (Mean+95%CI, - 11.59 (-7.1, -13.7)). UK-specific RBD substitutions (from -25.89 (-14.10, -32.5) to -38.72 (-26.70, - 49.80)) further strengthened antibody resistance (p< 0.001), as well as USA-specific RBD combinations (from -6.53 (-0.5, -4.6) to -18.11 (-7.6, -18.4)). Data of antibody neutralization to single RBD substitution was archived by liu, L. et al. Nature, 2021. Paired T-test was used for statistical analysis. (H) Wild type of SARS-CoV-2 RBD binds to ACE2 (PDB, 6M17). 15 WHO label RBD substitutions were displayed. (I-K) The common RBD combinations enhanced ACE2 binding accessibility and further enhanced by UK and USA specific RBD mutations. This analysis was conducted by Pymol mutagenesis.

### Specific RBD combinations enhanced antibody evasion and ACE2 binding accessibility

Supporting antibody evasion, recent evidence showed that the effect of individual RBD mutation to neutralization was largely diminished. However, the position adjacent RBD mutations may in fact synergistically contribute to antibody evasion. Here, we re-evaluate the synergistic effect of common and geographically specific RBD combinations on antibody evasion. Using the data of neutralization by 17 commercial monoclonal antibodies to single RBD mutation, we observed synergistic effects of common RBD combinations on antibody resistance (Mean+95%CI, -11.59 (-7.1, -13.7)) (Fig.1F). In addition, UK-specific mutations plus the common mutations significantly enhanced antibody evasion from -25.89 to -38.72 (p=0.0003). These USA-specific mutations plus the common mutations also enhanced antibody evasion from -6.53 to - 18.11 (p=0.0006).

Another profound impact of RBD combinations is ACE2 binding accessibility. Exploiting the recent released crystallography of spike protein and ACE2 complex, we evaluated RBD-ACE2 contact accessibility by mutagenesis (4) (Fig.1H). We primarily found that common RBD mutations with either UK-specific RBD mutations or USA-specific mutations largely increased ACE2 binding accessibility (Fig. 1I-K).

### Spatiotemporal transmission of Omicron variant with different RBD combinations

In the spike protein of Omicron variant, as mentioned, RBD mutations may concurrently exist and shows a geographical pattern of RBD combination. Therefore, its globally spatiotemporal transmission was tracked according to the pattern of RBD combinations and mutation linkage analysis. linkage disequilibrium (LD) analysis indicated that the geographical pattern of RBD combinations correlated with LD data. Of note, the six UK-specific RBD mutations can be further divided into two sub-groups of RBD combination (Fig. 2A). In line with this data, we found that geographical transmission and timely spread of Omicron can be well classified by the combinations of RBD mutations, and mainly dominated in continents of Europe and North America (Fig. 2B-C). Time-dependent frequency change suggested that the number of Omicron harboring two sub-combinations of UK-specific RBD mutations increased during the time of spillover, alongside with the decrease of Omicron harboring USA-specific RBD mutations (Fig.2D), especially after 2-Dec-2021 (Fig. 2E). Correlation analysis indicated that change of Omicron harboring the one of two UK RBD sub-combinations was positively related to the number of Omicron harboring another UK specific sub-combination, but was negatively related to the number of Omicron harboring USA specific RBD mutations (Fig. 2F-G). Collectively, combination of UK-specific RBD mutations essentially contribute to global transmission of SARS-CoV-2 Omicron variant, including sub-combination of S371L, S373P, and S375F, and sub-combination of Q498R, N501Y, and Y505H.

**Figure 2.**
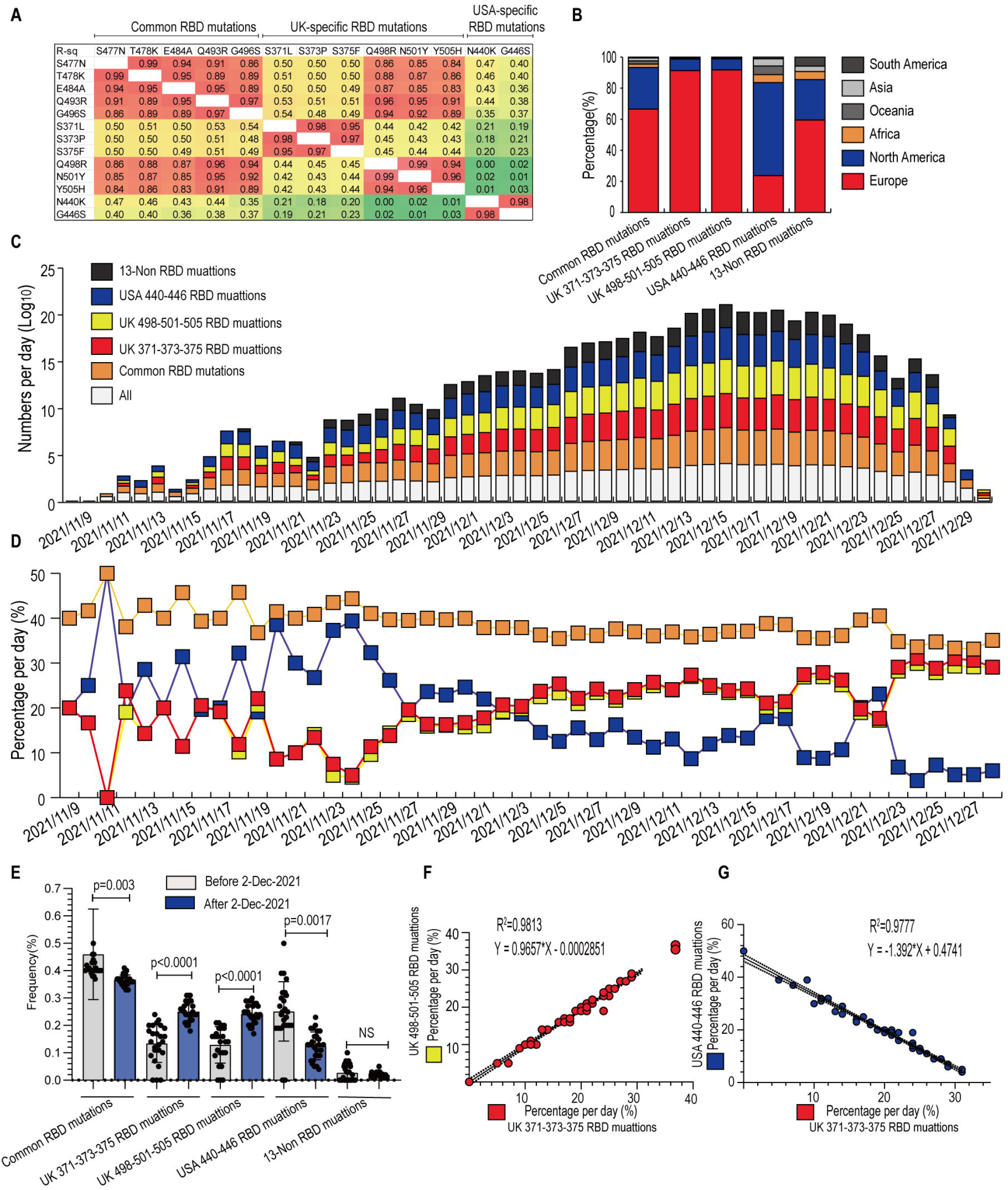
Spatiotemporal tracking of global Omicron variants with different RBD combinations. (A) linkage disequilibrium (LD) analysis indicated that the geographical pattern of RBD combinations correlated with LD data represented by R square. Of note, the six UK-specific RBD mutations can be further divided into two sub-groups of RBD combination. (B) Omicron with different RBD combinations has globally spread and can be well stratified by continents, in particular Europe and North America. (C) Time-dependent change suggested that global Omicron can be well classified by different RBD combination during the time of spillover. (D) However, the Omicron harboring the common, and USA-specific RBD mutations decreased along time, while the Omicron harboring either two sub-groups of UK-specific RBD mutations increased at the same period. (E) After 2-Dec-2021, the Omicron harboring USA-specific mutations was significantly decreased, while the Omicron with UK-specific mutations (UK-371-373-375 RBD mutations and UK-498-501-505 RBD mutations) was significantly increased. Student T test was used for statistical analysis. (F-G) Percentage per day of Omicron harboring UK-371-373-375 RBD mutations was positively related to the increased Omicron harboring UK-498-501-505 RBD mutations (R^2^=0.9813), but was negatively related to the decreased Omicron harboring USA-specific RBD mutations (R^2^=0.9777).

## Discussion

Currently, a large amount of spike protein mutations have been identified in Omicron variant, in which 15 out of 32 mutations located in RBD. As reported, these RBD mutations extensively escape neutralization of both monoclonal antibody or serum post COVID-19 vaccination. There are two mode-of-action of antibody evasion, either in form of single mutation or a combination of RBD mutations. A host of experimental proofs from multiple research centers have confirmed existence of antibody resistance, that is attribute to RBD mutations but in a form of single mutation or unknown combinations. As reported, single RBD mutation like K417N, G446S, E484A, S371L, N440K, G446S, and Q493R confers greater antibody resistance to Omicron variant (2, 3). In fact, the most possible mode-of-antibody resistance is combinations of RBD mutations. Supportively, we have identified that 5 out of 15 RBD mutations concurrently exist in both UK and USA Omicron. However, 6 out of 15 RBD mutations specifically exist in UK Omicron, and 2 RBD mutations in USA Omicron. This evidence indicated that RBD mutations existed in a manner of combination and showed geographical pattern. Compared with single RBD mutation, the RBD combinations largely escape antibody neutralization (fold change, -11.59) and even strengthened by UK or USA specific RBD mutations (fold change, -38.72, -18.11).

As antibody-binding RBD epitope overlaps with ACE2-binding motif. It is also possible that RBD combinations may change ACE2 binding affinity (4). Directed by this knowledge, structural variations of mutated RBD and ACE2 binding complex were analyzed. We overserved that the common RBD combinations enhanced ACE2 binding accessibility and was further enhanced by UK and USA specific RBD substitutions. Collectively, our data suggested that RBD combinations of Omicron existed and showed a patten of geographical combination. Spatiotemporal tracking of Omicron harboring different RBD combinations also showed geographical dominance and crossly transmitted between continents, especially in Europe and North America (6). Importantly, primary evidence from neutralization and ACE2 binding implied that RBD combinations and their geographical combinations largely intensified antibody evasion and enhanced ACE2 binding accessibility. Herein, by exploiting multiple geographical stratified GWAS studies, we have showed genetic evidence of RBD combinations. Despite significance of this knowledge, its biological impacts on antibody evasion and ACE2 binding were evaluated and confirmed by currently available experimental data. This new understanding will pave new avenue to study the exact impact of RBD combinations regarding antibody resistance and ACE2 binding.

## Materials and Methods

### GWAS study and LD analysis

The detailed sequencing data and methods are described in SI Appendix (6).

### Neutralization activity and ACE2 binding accessibility

The raw data of neutralization, data processing and statistic are described in SI Appendix.

### Data availability

Raw data and processed CSV files are available at supplementary dataset or with contacts to corresponding author for details.

## Acknowledgments

This work was supported by National Natural Science Foundation of China (32102706 to X. Ou), Sichuan Veterinary Medicine and Drug Innovation Group of China Agricultural Research System (SCCXTD-2021-18), Science and Technology Program of Sichuan Province (2020YJ0396 to X. Ou), China Agriculture Research System of MOF and MARA.

## References

1. WHO (2021) Classification of Omicron (B.1.1.529): SARS-CoV-2 Variant of Concern.

2. L. Liu et al., Striking Antibody Evasion Manifested by the Omicron Variant of SARS-CoV-2. Nature10.1038/s41586-021-04388-0 (2021).

3. D. Planas et al., Considerable escape of SARS-CoV-2 Omicron to antibody neutralization. Nature 10.1038/s41586-021-04389-z (2021).

4. R. H. Yan et al., Structural basis for the recognition of SARS-CoV-2 by full-length human ACE2. Science 367, 1444-+ (2020).

5. X. Ou et al., Tracing two causative SNPs reveals SARS-CoV-2 transmission in North America population. BioRvix https://doi.org/10.1101/2020.05.12.092056 (2020).

6. X. M. Ou et al., Tracing genetic signatures of bat-to-human coronaviruses and early transmission of North American SARS-CoV-2. Transbound Emerg Dis 10.1111/tbed.14148 (2021).

