## Supplementary file for "Microbial GWAS studies revealing combinations of Omicron RBD mutations existed and may contribute to antibody evasion and ACE2 binding"

**This PDF file includes:**

Supplementary text

Legends for Datasets S1 to S6

SI References

Supplementary Information Text

**Extended methods**

**Data acquisition.**

The human SARS-CoV-2 isolated from Wuhan, China was achieved from the NCBI database (GenBank No.: MN908947.3). For GWAS study, full-genome omicron sequences of global SARS-CoV-2 collected from 5-Nov-2021 to 02-Jan-2021 (12:00 GMT + 8) were archived from the database of GISAID Initiative EpiCoV platform (GISAID; <https://www.epicov.org>). A total of 100857 omicron sequences were archived and filtered by the criteria, including high coverage only (> 29, 000 bp, 1X coverage of genome), exclusion of low coverage, and the sequence with unconfident bases (N) inside. The identical sequences were further removed by CD-HIT software (version 4.8.1, parameters: -aL 1 -aS 1 -c 1 -s 1) (1). A final 77123 sequences were used in this study.

**GWAS and linkage disequilibrium (LD) analysis.**

The SNPs and INDELs polymorphisms were detected by MUMmer software (version 3.0, nucmer, show-snps) (2) using the Wuhan-Hu-1 strain (GISAID: EPI_ISL_402125, Genbank: NC_045512.2) as a reference genome. To identify geographic associated SNPs in the population of South Africa, United Kingdom and United State, three geographic stratified genome-wide association studies (GWASs) were performed using PLINK software (version 1.90) (3). The details of GWAS study can refer our previous study (4).The empirical threshold of p-value was calculated according to Benjamini & Hochberg method (1995) (5). The top SNPs were listed (Dataset S1-4) and the LD of pairing SNPs were estimated by Haploview software (version 4.1) (6).

**Evaluating antibody neutralization of RBD combinations.**

Antibody resistance to 17 commercial mono-antibody was evaluated by combinations of RBD substitution. Antibody resistance to UK-specific and USA-specific RBD combinations were also estimated. Raw data of antibody neutralization to single RBD substitution was archived by liu, L. et al. Nature, 2021(Dataset S5). Paired T-test was used for statistical analysis.

**Spatiotemporal evolution of global Omicron variant.**

To analyze the accumulation trend of RBD combinations after transmitting from South Africa, the frequencies of omicron variants with 5 common RBD mutations per day were counted, as well as these variants with UK or USA-specific mutations. These analyses were performed by Microsoft® Excel 2016.

**Statistical analysis.**

Data from the section of SNPs accumulating analysis were plotted by Graphpad (Version 8.2.1). Paired T test was used for statistical significance of antibody neutralization. P values less than 0.05 are considered significant.

Dataset S1. Common SNPs identified by three geographic stratified GWAS studies (SA(n=1383), UK (n=39849) and USA (n=19534)).

Dataset S2. UK-specific SNPs identified by SA (n=1383) and UK (n=39849) GWAS study.

Dataset S3. USA-specific SNPs identified by SA (n=1383) and USA (n=19534) GWAS study.

Dataset S4. RBD combinations in UK and USA Omicron.

Dataset S5. Raw data of antibody neutralization to RBD combinations.

Dataset S6. LD analysis of RBD mutations of Omicron variant.

**SI References**

1.	Y. Huang, B. Niu, Y. Gao, L. Fu, W. Li, CD-HIT Suite: a web server for clustering and comparing biological sequences. *Bioinformatics* **26**, 680-682 (2010).

2.	S. Kurtz *et al.*, Versatile and open software for comparing large genomes. *Genome biology* **5**, R12 (2004).

3.	S. Purcell *et al.*, PLINK: a tool set for whole-genome association and population-based linkage analyses. *The American journal of human genetics* **81**, 559-575 (2007).

4.	X. M. Ou *et al.*, Tracing genetic signatures of bat-to-human coronaviruses and early transmission of North American SARS-CoV-2. *Transbound Emerg Dis* 10.1111/tbed.14148 (2021).

5.	Y. Benjamini, Y. Hochberg, Controlling the false discovery rate: a practical and powerful approach to multiple testing. *Journal of the Royal statistical society: series B* **57**, 289-300 (1995).

6.	J. C. Barrett, B. Fry, J. Maller, M. J. Daly, Haploview: analysis and visualization of LD and haplotype maps. *Bioinformatics* **21**, 263-265 (2005).
